# Estimating COVID-19 Vaccination Effectiveness Using Electronic Health Records of an Academic Medical Center in Michigan

**DOI:** 10.1101/2022.01.29.22269971

**Authors:** Emily K. Roberts, Tian Gu, Bhramar Mukherjee, Lars G. Fritsche

## Abstract

**Importance:** Systematic characterization of the protective effect of vaccinations across time and at-risk populations is needed to inform public health guidelines and personalized interventions.

**Objective:** To evaluate the vaccine effectiveness (VE) over time and determine differences across demographic and clinical risk factors of COVID-19.

**Design, Setting, and Participants:** This test negative design consisted of adult patients who were tested or diagnosed for COVID-19 at Michigan Medicine in 2021. Variables extracted from Electronic Health Records included vaccination status, age, gender, race/ethnicity, comorbidities, body mass index, residential-level socioeconomic characteristics, past COVID-19 infection, being immunosuppressed, and health care worker status.

**Exposure:** The primary exposure was vaccination status and was categorized into fully vaccinated with and without booster, partially vaccinated, or unvaccinated.

**Main Outcomes and Measures:** The main outcomes were infection with COVID-19 (positive test or diagnosis) and having severe COVID-19, i.e., either being hospitalized or deceased. Based on these, VE was calculated by quarter, vaccine, and patient characteristics.

**Results:** Of 170,487 COVID-19 positive adult patients, 78,002 (45.8%) were unvaccinated, and 92,485 (54.2%) were vaccinated, among which 74,060 (80.1%) were fully vaccinated. COVID-19 positivity and severity rates were substantially higher among unvaccinated (12.1% and 1.4%, respectively) compared to fully vaccinated individuals (4.7% and 0.4%, respectively). Among 7,187 individuals with a booster, only 18 (0.3%) had a severe outcome. The covariate-adjusted VE against an infection was 62.1% (95%CI 60.3–63.8%), being highest in the Q2 of 2021 (90.9% [89.5–92.1%]), lowest in Q3 (60.1% [55.9–64.0%]), and rebounding in Q4 to 68.8% [66.3– 71.1%]). Similarly, VE against severe COVID-19 overall was 73.7% (69.6–77.3%) and remained high throughout 2021: 87.4% (58.1–96.3%), 92.2% (88.3–94.8%), 74.4% (64.8–81.5%) and 83.0% (78.8–86.4%), respectively. Data on fully vaccinated individuals from Q4 indicated additional protection against infection with an additional booster dose (VE-Susceptibility: 64.0% [61.1–66.7%] vs. 87.3% [85.0–89.2%]) and severe outcomes (VE-Severity: 78.8% [73.5–83.0%] vs. 94.0% [89.5–96.6%]). Comparisons between Pfizer-BioNTech and Moderna vaccines indicated similar protection against susceptibility (82.9% [80.7–84.9%] versus 88.1% [85.5– 90.2%]) and severity (87.1% [80.3–91.6%]) vs. (84.9% [76.2–90.5%]) after controlling for vaccination timing and other factors. There was no significant effect modification by all the factors we examined.

**Conclusions and Relevance:** Our findings suggest that COVID-19 vaccines offered high protection against infection and severe COVID-19, and showed decreasing effectiveness over time and improved protection with a booster.

**Key Points:** *Question:* How do the rates of COVID-19 outcomes (infections or mild/severe disease) compare across vaccination status and quarters of 2021, after adjusting for confounders?

*Findings:* In this cohort of 170,487 adult patients tested for or diagnosed with COVID-19 during 2021, both COVID-19 positivity and severity rates were substantially higher in unvaccinated compared to fully vaccinated individuals. Vaccine effectiveness estimation was adjusted for covariates potentially related to both being vaccinated and COVID-19 outcomes; this also allowed us to determine if effectiveness differed across patient subgroups. The estimated vaccine effectiveness across the four quarters of 2021 was 62.1% against infection and was 73.7% against severe COVID-19 (defined as hospitalization, ICU admission, or death). There was no significant effect modification by all the factors we examined.

*Meaning:* These findings suggest COVID-19 vaccines had relatively high protection against infection and severe COVID-19 during 2021 for those who received two doses of an mRNA vaccine (Moderna or Pfizer-BioNTech) or one dose of the Janssen vaccine, of which the effectiveness decreased over time and improved with a booster.

## Introduction

Three Coronavirus Disease 2019 (COVID-19) vaccines were developed, assessed for efficacy against symptomatic COVID-19 disease in placebo-controlled trials, and approved under emergency use authorization in the United States by February 2021: mRNA-1273 (Moderna), BNT162b2 (Pfizer-BioNTech), and Ad26.COV2.S (Johnson & Johnson-Janssen). As of January 26, 2022, 536 million doses have been administered, and 210 million people are fully vaccinated, meaning they received at least one dose of Janssen or two doses of Pfizer-BioNTech or Moderna vaccine, in the United States^1^. In light of new SARS-CoV-2 variants, observational studies can provide insight on vaccine effectiveness (VE) with respect to severity of disease and measures of effectiveness over time as a proxy for protection against different variants.

Other studies have examined the effectiveness of the vaccines in real world settings over shorter and earlier periods of time and concluded the VE is relatively high against positivity and disease severity such as hospitalization and death^2-9^. While these studies support evidence of robust protection against variants with modest waning of VE through the third quarter of 2021, the FDA approved booster shots for continued protection of the vaccinated population over time. Still, the time-varying protection of vaccination over the course of 2021 and effectiveness among different vaccine brands remain unclear. More data from healthcare centers is needed to track hospitalizations and deaths across vaccination status. It is important to adjust for covariates that may be related to both getting vaccinated and severe COVID-19 outcomes as potential confounders in real-world studies^10^. Electronic health records (EHRs) are helpful for this type of research as they allow for such big data and covariate adjustment.

The objective of this work is to evaluate VE and determine if any patient characteristics show effect modification of VE using EHR from the University of Michigan Health System, in short Michigan Medicine (MM), a nationally ranked healthcare center and one of the largest health care complexes in the state of Michigan. Specifically, this time-stratified retrospective cohort study has two goals: (i) to investigate the COVID-19 VE against test positivity and severe COVID-19 outcomes across 2021, and (ii) to examine VE stratified by the two most common vaccines Pfizer-BioNTech and Moderna, and by sociodemographic and clinical characteristics that are associated with COVID-19 outcomes.

## Methods

### Study cohort and COVID-19 vaccination

Our study sample using the test negative design consisted of 170,487 adult patients who were tested or diagnosed for COVID-19 at MM during the four quarters of 2021, including 14,059 tested positive / diagnosed and 156,428 tested negative. Tested patients received a total 963,726 tests of the six different types: an in-house polymerase chain reaction (PCR) test (336,631 tests (34.9%)), a point of care PCR test (58,644 tests (6.1%)), a commercial PCR test (Viracor; 467 tests (0.048%); Quest 37 tests (0.004%)), COVID-19 nasopharynx or oropharynx PCR tests deployed by the Michigan Department of Health and Human Services (71 tests (0.007%)), and a small fraction of ribonucleic acid (RNA) tests (seven test (0.001%)); 567,869 test results (58.9%) were from patients transferred, tested elsewhere, or had no information on the test type.

Among all patients, 78,002 had no documented vaccination (“unvaccinated/unknown”) and 92,485 were vaccinated. Among those vaccinated, 74,060 were fully vaccinated according to FDA’s vaccination guideline, meaning they received either two doses of Moderna or Pfizer-BioNTech, or one dose of Janssen vaccine, and least 21 days had passed since completion of the series. The remaining 18,425 vaccinated patients either had not completed their vaccination series or did not adhere to the guidelines and received a mixed sequence of the above-mentioned vaccines (**Table 1, Table S1**). 254 individuals who received unspecified or other brand vaccines (AstraZeneca, Sinopharm, Sinovac, or Novavac) were not included in the study sample. Being boosted was defined as receiving any additional vaccination any time after the original full vaccination schedule.

**Table 1.** Characteristics of the Michigan Medicine cohort of 170,487 individuals who were tested for or diagnosed with COVID-19 between January and December 2021. For individuals who were tested across multiple quarters, only the vaccination status at their last observed quarter is shown.

|  | Vaccination Status |  |  |  |
| --- | --- | --- | --- | --- |
|  | Unvaccinated / Unknown | Partially Vaccinated | Fully Vaccinated* | Fully Vaccinated & ≥ 1 Booster |
| n | 78002 | 18425 | 74060 | 7187 |
| Age, mean (SD) | 46.9 (19) | 53.7 (19.4) | 55 (19.7) | 60 (18.2) |
| median (IQR) | 46 (31.8) | 56.4 (31.3) | 58.2 (32.8) | 64.5 (26) |
| Age Bin, n (%) |  |  |  |  |
| 18 – 49.9 | 43430 (55.7) | 7286 (39.5) | 28130 (38) | 1981 (27.6) |
| 50 – 64.9 | 18986 (24.3) | 5127 (27.8) | 18094 (24.4) | 1670 (23.2) |
| >= 65 | 15586 (20) | 6012 (32.6) | 27836 (37.6) | 3536 (49.2) |
| Male Gender, n (%) | 33023 (42.3) | 7804 (42.4) | 30924 (41.8) | 3122 (43.4) |
| BMI, mean (SD) | 29.2 (7.6) | 29.1 (7.2) | 29 (7) | 28.9 (6.6) |
| BMI Bin, n (%) |  |  |  |  |
| < 18.5 | 20007 (29.7) | 4798 (29.1) | 20063 (29.7) | 2044 (30) |
| 18.5 – 24.9 | 1497 (2.2) | 280 (1.7) | 1136 (1.7) | 94 (1.4) |
| 25 – 29.9 | 20027 (29.8) | 5220 (31.6) | 21348 (31.7) | 2210 (32.4) |
| >= 30 | 25777 (38.3) | 6206 (37.6) | 24894 (36.9) | 2470 (36.2) |
| Race Ethnicity, n (%) |  |  |  |  |
| Caucasian / Non-Hispanic | 55111 (70.7) | 13853 (75.2) | 56883 (76.8) | 5830 (81.1) |
| African American / Non-Hispanic | 9187 (11.8) | 1593 (8.6) | 5287 (7.1) | 349 (4.9) |
| Other / Non-Hispanic or Hispanic | 6918 (8.9) | 1773 (9.6) | 7808 (10.5) | 673 (9.4) |
| Other / Unknown Ethnicity | 6786 (8.7) | 1206 (6.5) | 4082 (5.5) | 335 (4.7) |
| Primary Care at MM, n (%) | 24847 (31.9) | 7074 (38.4) | 33192 (44.8) | 3697 (51.4) |
| NDI without Proportion Black, n (%) |  |  |  |  |
| Quartile 1 | 22022 (34.9) | 6684 (42.4) | 27826 (45.4) | 3116 (50.9) |
| Quartile 2 | 14860 (23.6) | 3729 (23.6) | 14545 (23.7) | 1359 (22.2) |
| Quartile 3 | 13808 (21.9) | 3167 (20.1) | 11920 (19.4) | 1128 (18.4) |
| Quartile 4 | 12390 (19.6) | 2197 (13.9) | 7038 (11.5) | 515 (8.4) |
| Persons Per Square Mile, n (%) |  |  |  |  |
| Quartile 1 | 18615 (29.5) | 4166 (26.4) | 15058 (24.6) | 1340 (21.9) |
| Quartile 2 | 18826 (29.8) | 5074 (32.2) | 20076 (32.7) | 2139 (35) |
| Quartile 3 | 19742 (31.3) | 5128 (32.5) | 20778 (33.9) | 2107 (34.4) |
| Quartile 4 | 5897 (9.3) | 1409 (8.9) | 5417 (8.8) | 532 (8.7) |
| Elixhauser Score AHRQ, mean (SD) | 3.6 (10.7) | 4.9 (11.6) | 5.9 (12.5) | 8.9 (14.2) |
| Elixhauser Score AHRQ Bin, n (%) |  |  |  |  |
| <0 | 19887 (30) | 4752 (28.7) | 18374 (27.4) | 1543 (22.8) |
| 0 | 18177 (27.4) | 3900 (23.5) | 15036 (22.4) | 1135 (16.8) |
| 1-4 | 7324 (11.1) | 1937 (11.7) | 7036 (10.5) | 706 (10.5) |
| >=5 | 20837 (31.5) | 5988 (36.1) | 26624 (39.7) | 3370 (49.9) |
| Past COVID-19 Infection, n (%) | 1728 (2.2) | 381 (2.1) | 1401 (1.9) | 79 (1.1) |
| Health Care Worker, n (%) | 571 (0.7) | 233 (1.3) | 2786 (3.8) | 465 (6.5) |
| Past Immunosuppression, n (%) | 3062 (3.9) | 962 (5.2) | 4331 (5.8) | 818 (11.4) |

\* Includes individuals who received booster

### COVID-19 outcomes

In this study, we focused on two COVID-19 associated outcomes: susceptibility, i.e., tested positive / diagnosed for COVID-19, and severity, i.e., having severe COVID-19 defined as either being hospitalized, admitted to an intensive care unit (ICU) within one month after a positive COVID-19 test / diagnosis, or died within two months after a positive COVID-19 test / diagnosis (**Table 2**). The summary of these outcomes is listed in Table 2, stratified by vaccination status.

**Table 2.**
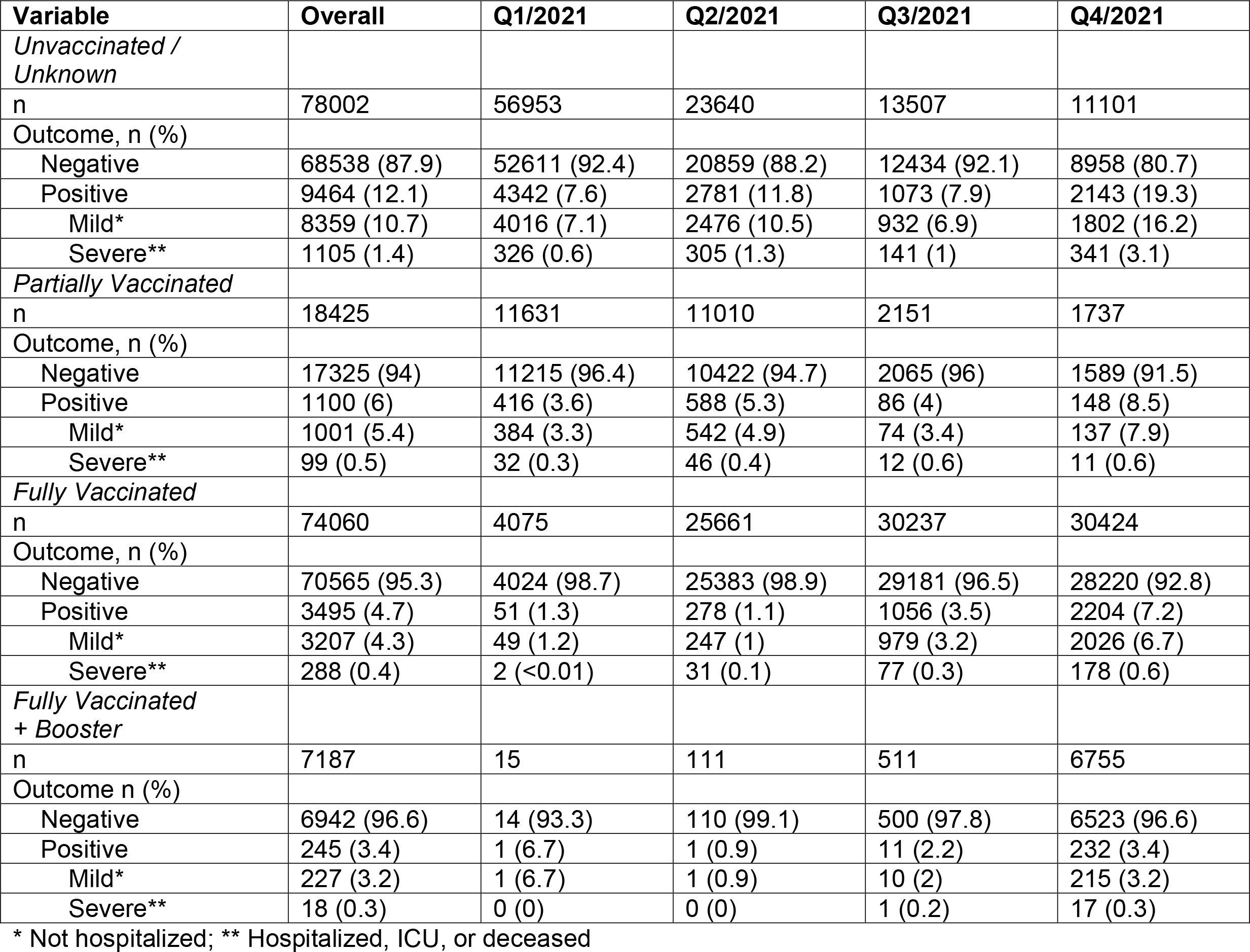
COVID-19 Outcome Summary of 170,487 individuals who were tested for or diagnosed with COVID-19 between January and December 2021. For individuals who were tested/diagnosed across multiple quarters, the vaccination status at their last observed quarter is summarized in the overall cohort.

| **Variable** | **Overall** | **Q1/2021** | **Q2/2021** | **Q3/2021** | **Q4/2021** |
| --- | --- | --- | --- | --- | --- |
| *Unvaccinated / Unknown* |  |  |  |  |  |
| n | 78002 | 56953 | 23640 | 13507 | 11101 |
| Outcome, n (%) |  |  |  |  |  |
| Negative | 68538 (87.9) | 52611 (92.4) | 20859 (88.2) | 12434 (92.1) | 8958 (80.7) |
| Positive | 9464 (12.1) | 4342 (7.6) | 2781 (11.8) | 1073 (7.9) | 2143 (19.3) |
| Mild* | 8359 (10.7) | 4016 (7.1) | 2476 (10.5) | 932 (6.9) | 1802 (16.2) |
| Severe** | 1105 (1.4) | 326 (0.6) | 305 (1.3) | 141 (1) | 341 (3.1) |
| *Partially Vaccinated* |  |  |  |  |  |
| n | 18425 | 11631 | 11010 | 2151 | 1737 |
| Outcome, n (%) |  |  |  |  |  |
| Negative | 17325 (94) | 11215 (96.4) | 10422 (94.7) | 2065 (96) | 1589 (91.5) |
| Positive | 1100 (6) | 416 (3.6) | 588 (5.3) | 86 (4) | 148 (8.5) |
| Mild* | 1001 (5.4) | 384 (3.3) | 542 (4.9) | 74 (3.4) | 137 (7.9) |
| Severe** | 99 (0.5) | 32 (0.3) | 46 (0.4) | 12 (0.6) | 11 (0.6) |
| *Fully Vaccinated* |  |  |  |  |  |
| n | 74060 | 4075 | 25661 | 30237 | 30424 |
| Outcome, n (%) |  |  |  |  |  |
| Negative | 70565 (95.3) | 4024 (98.7) | 25383 (98.9) | 29181 (96.5) | 28220 (92.8) |
| Positive | 3495 (4.7) | 51 (1.3) | 278 (1.1) | 1056 (3.5) | 2204 (7.2) |
| Mild* | 3207 (4.3) | 49 (1.2) | 247 (1) | 979 (3.2) | 2026 (6.7) |
| Severe** | 288 (0.4) | 2 (<0.01) | 31 (0.1) | 77 (0.3) | 178 (0.6) |
| *Fully Vaccinated + Booster* |  |  |  |  |  |
| n | 7187 | 15 | 111 | 511 | 6755 |
| Outcome n (%) |  |  |  |  |  |
| Negative | 6942 (96.6) | 14 (93.3) | 110 (99.1) | 500 (97.8) | 6523 (96.6) |
| Positive | 245 (3.4) | 1 (6.7) | 1 (0.9) | 11 (2.2) | 232 (3.4) |
| Mild* | 227 (3.2) | 1 (6.7) | 1 (0.9) | 10 (2) | 215 (3.2) |
| Severe** | 18 (0.3) | 0 (0) | 0 (0) | 1 (0.2) | 17 (0.3) |

\* Not hospitalized; \*\* Hospitalized, ICU, or deceased

### Definition of demographics, socioeconomic status, and other covariates

We extracted the EHR data from patients with COVID-19 diagnoses or test results for COVID-19 (positive or negative) at MM, including age, self-reported gender, self-reported race/ethnicity, body mass index (BMI; calculated as weight in kilograms divided by height in meters squared), Neighborhood Socioeconomic Disadvantage Index (NDI) without proportion of Black (coded as quartiles, with larger quartiles representing more disadvantaged communities), and population density measured in persons per square mile (coded as quartiles). The Elixhauser comorbidity score developed by the Agency for Healthcare Research and Quality (AHRQ) was calculated to comprehensively characterize patients’ pre-existing comorbidity conditions using ICD9 and ICD10 codes and the R package “comorbidity” ^11^. Past COVID-19 infection was defined as testing positive at least 21 days before the first test of a quarter (if a person remained negative in that quarter), or 21 days before the first positive test of a quarter (if tested positive at least once in that quarter). Being immunosuppressed was defined for patients who had any immunosuppressants listed in their medication or prescription records within one year before a corresponding test (Calcineurin inhibitors, Interleukin inhibitors, TNF-alfa inhibitors, or any selective or other immunosuppressants)^12^. In addition, health care worker status was defined based on documented participation in a Health Care Worker Screen or a SARS-CoV-2 PCR test order for Health Care Workers. We report the variables’ missingness by vaccination status in Table S2. Missing data were handled by using a complete case analysis.

### Statistical analysis

We fit Firth’s bias-corrected logistic regression for each COVID-19 related outcome *Y*_*COVID*_, considering several sets of covariates:

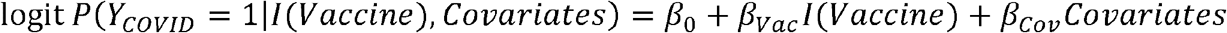

where 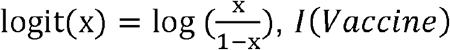 is the indicator of vaccination status, either with or without a booster, i.e. 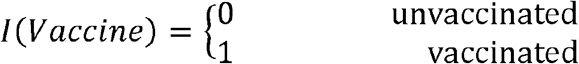 for them models comparing fully vaccinated individuals, and

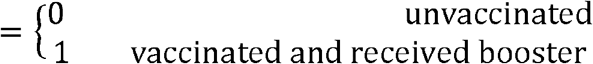

for the models comparing boosted individuals, and *Covariates* denotes the vector of covariates adjusted. Five nested sets of *Covariates* were evaluated, denoted as adjustment 1-5 (defined in **Table S3**). We reported the final model adjusted for age, gender, race/ethnicity, Elixhauser score, NDI without proportion of Black, population density, past COVID-19 infection, and health care worker status (adjustment 5).

We estimated the vaccine efficacy (VE) through the following formula ^13^:

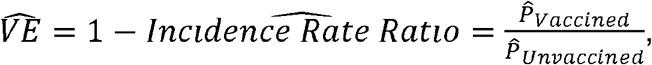

where *Incidence Rate Ratio*, the ratio of incidence between the vaccinated group and the unvaccinated group, can be estimated using the Michigan 12-month COVID-19 incidence rate (IR) and the estimated odds ratio (OR) from above logistic regression through the following formula ^14^:

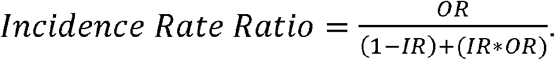

We estimated an IR of 0.088 based on the cumulative case count until December 29, 2021 (1,710,325 cumulative cases; JHU COVID-19 Dashboard ^15^ and the resident population of Michigan (Census 2020: 10,077,331) ^16^.

In addition, we conducted a quarter-stratified analysis to examine the time-varying changes in vaccination effectiveness over 2021: Quarter 1 (Q1), January 1, 2021–March 31, 2021; Quarter 2 (Q2), April 1, 2021–June 30, 2021; Quarter 3 (Q3), July 1, 2021–September 30, 2021; Quarter 4 (Q4), October 1, 2021–December 31, 2021.

To assess the potentially different VE across different strata of risk factors (denoted as *X*) associated with COVID-19 outcomes, we further conducted interaction analysis by vaccine status using the following model:

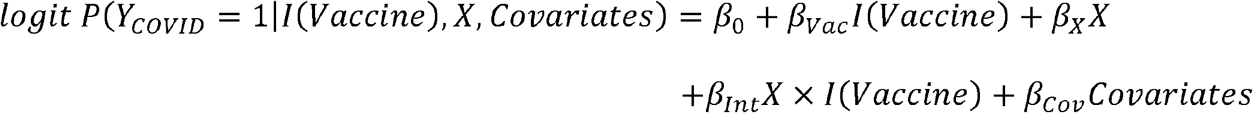

All analyses were performed in R statistical software version 4.1.2 (R Project for Statistical Computing). For each model, we reported VE along with the corresponding 95% Wald confidence intervals (CI) and *P* values. For the interaction analysis, we obtained the subgroup OR based on *β*_*vac*_ + *β*_*Int*_ and reported the subgroup VE along with the *P* values, *P*_*int*_, by testing the difference of subgroup effects via the null hypothesis *H*_0_ : *β*_*int*_ = 0.

## Results

### Study characteristics

In our cohort of 170,487 adults, 92,485 (54.2%) had at least one COVID-19 vaccination dose before their test. The remaining 78,002 individuals had no documented vaccination record in their EHR and thus were considered unvaccinated though they may include individuals who received their vaccination elsewhere. A total of 74,060 had a complete vaccination series (“fully vaccinated”) and 18,425 had an incomplete vaccination series or a mix of vaccines, e.g., Moderna – Pfizer – Moderna (**Table 1, Table S1**).

We observed that mean age, Elixhauser Score, and proportion of health care workers was lowest in unvaccinated and highest in the fully vaccinated groups. The proportion of individuals who received Primary Care at Michigan was higher in the partially and fully vaccinated groups compared to the unvaccinated/unknown subset (38.4% and 44.8% versus 31.9%). Among those fully vaccinated, the 7,187 boosted individuals were characterized by the highest mean Elixhauser Score (8.9 versus <= 5.9) and an enrichment of individuals who received immunosuppressants in the past (11.4% versus <= 5.4%) (**Table 1**).

### Observed Outcomes

When assessing COVID-19 positivity in the overall cohort, we observed substantially higher rates in unvaccinated (12.1%) compared to partially vaccinated (6.0%) or fully vaccinated individuals (4.7%). The positivity rate among fully vaccinated individuals reached a valley with low community transmission in Q2 (Q1: 1.3% and Q2: 1.1%) and surged to 3.2% in Q3 and to 6.7% in Q4 (**Table 2**) indicating potential waning effectiveness or consequences of spreading of the more immune evasive SARS-CoV-2 B.1.617.2 (Delta) and the B.1.1.159 (Omicron) variants^17-19^.

The overall rate of severe COVID-19 (hospitalization and/or ICU stay or death) was highest in unvaccinated (1.4%) compared to partially (0.5%) and fully vaccinated individuals (0.4%). Across the four quarters of 2021, the severity rate among fully vaccinated individuals remained relatively low (Q1: <0.01%, Q2: 0.1%, Q3: 0.3%, and Q4: 0.6%) compared to the unvaccinated group (Q1: 0.6%, Q2: 1.3%, Q3: 1.0%, Q4: 3.1%) indicating a less strong waning effect of the vaccine-mediated protection against severe COVID-19. Only 18 severe cases (0.3%) were observed among the 7,187 fully vaccinated individuals who received a booster shot (**Table 2**).

### Vaccine Effectiveness

To next estimate the VE of a full vaccination against an infection (VE-Susceptibility) and against severe COVID-19 (VE-Severity), we applied logistic regression models unadjusted and adjusted for relevant covariates (see **Methods, Table S3**). To reduce bias in the estimates, it is important to adjust for potential confounding factors that may be related to both getting vaccinated (our exposure) and outcome (COVID positivity and severity) which include age, gender, race/ethnicity, comorbidity score, residential-level socioeconomic characteristics, past COVID-19 Infection, and health care worker status. In **Figure 1**, we present VE estimates for the overall and the time-stratified cohort (Q1, Q2, Q3, and Q4). Since the VE estimates were reasonably consistent across adjustments, e.g., in the overall cohort VE-Susceptibility ranged between 59.4 and 62.1%, we will only discuss the estimates from the most conservative setting (Adjustment 5 which included all above mentioned covariates) in the following. Results from all models are included in the corresponding figures for the sake of completeness. VE-Susceptibility in Q1 was 82.0% (76.0–86.5%), increased in Q2 to 90.9% (89.5–92.1%), and was substantially reduced in Q3 (60.1% [55.9-64.0%]) and Q4 (68.8% [66.3–71.1%]), suggesting waning vaccine effectiveness over time and potentially improved protection due to the increasing number of individuals who received booster shots (**Figure 1A**). This pattern was also observed for VE-Severity estimates though they were generally higher and did not drop off as dramatically (Q1: 87.4% [58.1–96.3%], Q2: 92.2% [88.3–94.8%], Q3: 74.4% [64.8–81.5%], Q4: 83.0% [78.8–86.4%]; **Figure 1B**).

**Figure 1.**
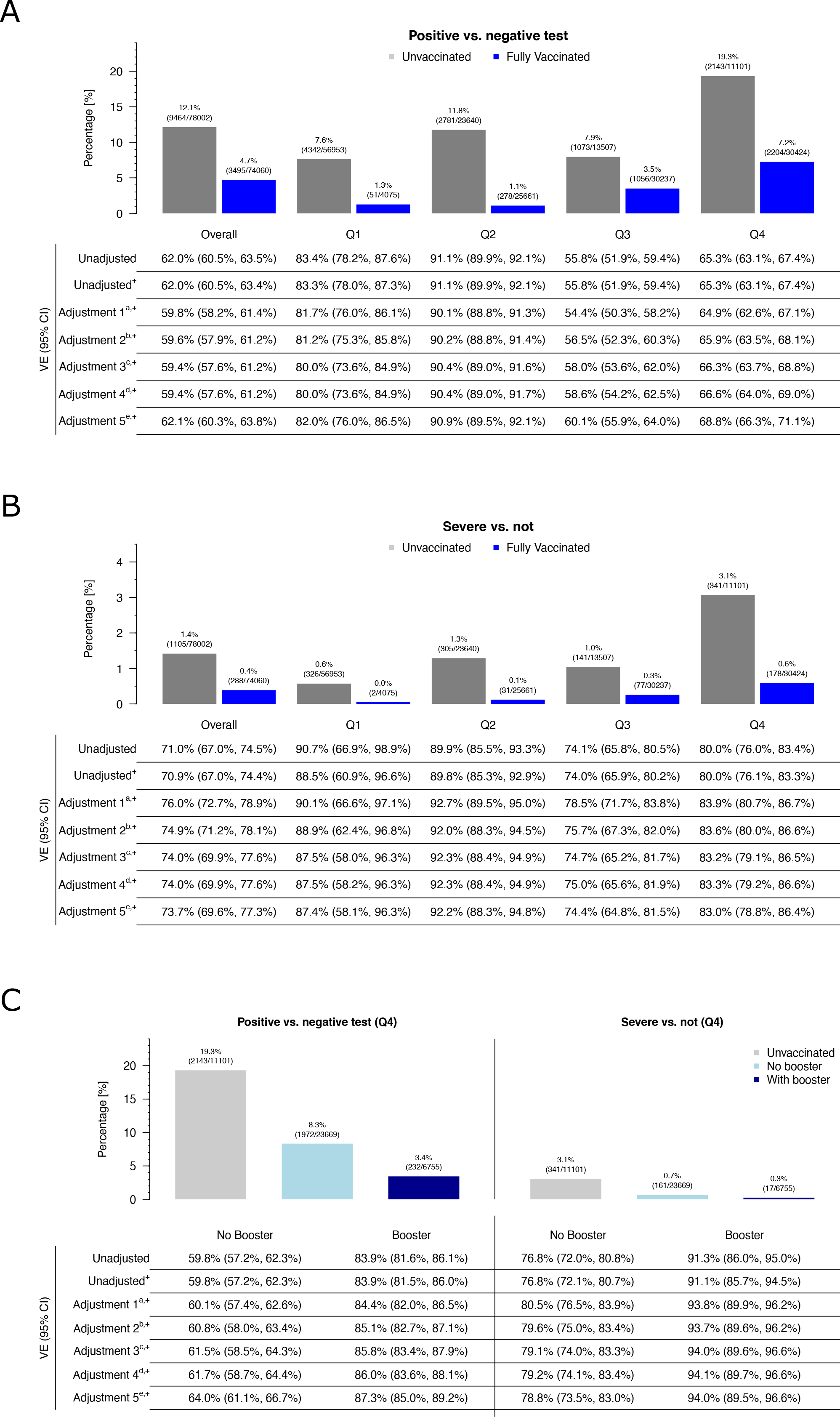
Vaccine Effectiveness in 2021 regarding (A) Positivity and (B) Severity, as well as a comparison between fully vaccinated without or with booster in 2021/Q4 (C). Notes: +: Logistic regression; Adjustment 1: Age, Gender, Race/Ethnicity; Adjustment 2: Adjustment 1 + ElixhauserScore_AHRQ; Adjustment 3: Adjustment 2 + PersonsPerSquareMile + NDIwithoutProportionBlack; Adjustment 4: Adjustment 3 + Past COVID-19 Infection; Adjustment 5: Adjustment 4 + HealthCareWorkerStatus

Considering the increase in vaccine booster shots around Q4, we conducted a stratified analysis by booster status. These results of VE in Q4 demonstrated additional protection of a booster against infection (VE-Susceptibility: no booster: 64.0% [61.1–66.7%] vs. booster: 87.3% [85.0–89.2%]) and severe outcomes (VE-Severity: no booster: 78.8% [73.5–83.0%] vs. booster: 94.0% [89.5–96.6%]; **Figure 1C, Figure S1)**.

We also performed a sensitivity analysis using only data on individuals who received primary care at Michigan Medicine and thus were more likely have fully documented vaccination data. In this analysis, all estimates were substantially higher for VE-Susceptibility (Q1: 88.7% [81.5–93.1%], Q2: 93.8% [92.3–95.0%], Q3: 68.7% [63.7–73.0%], Q4: 72.3 [68.9–75.4%]**; Figure S2A**) and VE-Severity (Q1: not available / no severe case among fully vaccinated, Q2: 95.2% [91.1–97.4%], Q3: 79.4% [68.0–86.7%], Q4: 80.8% [73.8–85.9%]; **Figure S2B**) indicating that the VE estimates in the full cohort might be influenced by incomplete vaccination documentation in the EHR (**Figure S2**).

### Comparing VE between Pfizer-BioNTech and Moderna

Since the majority of the 74,060 fully vaccinated individuals completed a vaccine series with either Pfizer-BioNTech (n = 45,168, 61.0%) or Moderna (n = 25,267, 34.1%), we next compared the effectiveness of the two vaccines. To reduce the differing impact of a waning effectiveness, i.e., the Pfizer-BioNTech vaccine was available earlier than the Moderna vaccine, we focus on the analysis which only included individuals who reached full vaccination status less than 3 months before their COVID-19 test or diagnosis (**Figure 2**).

**Figure 2.**
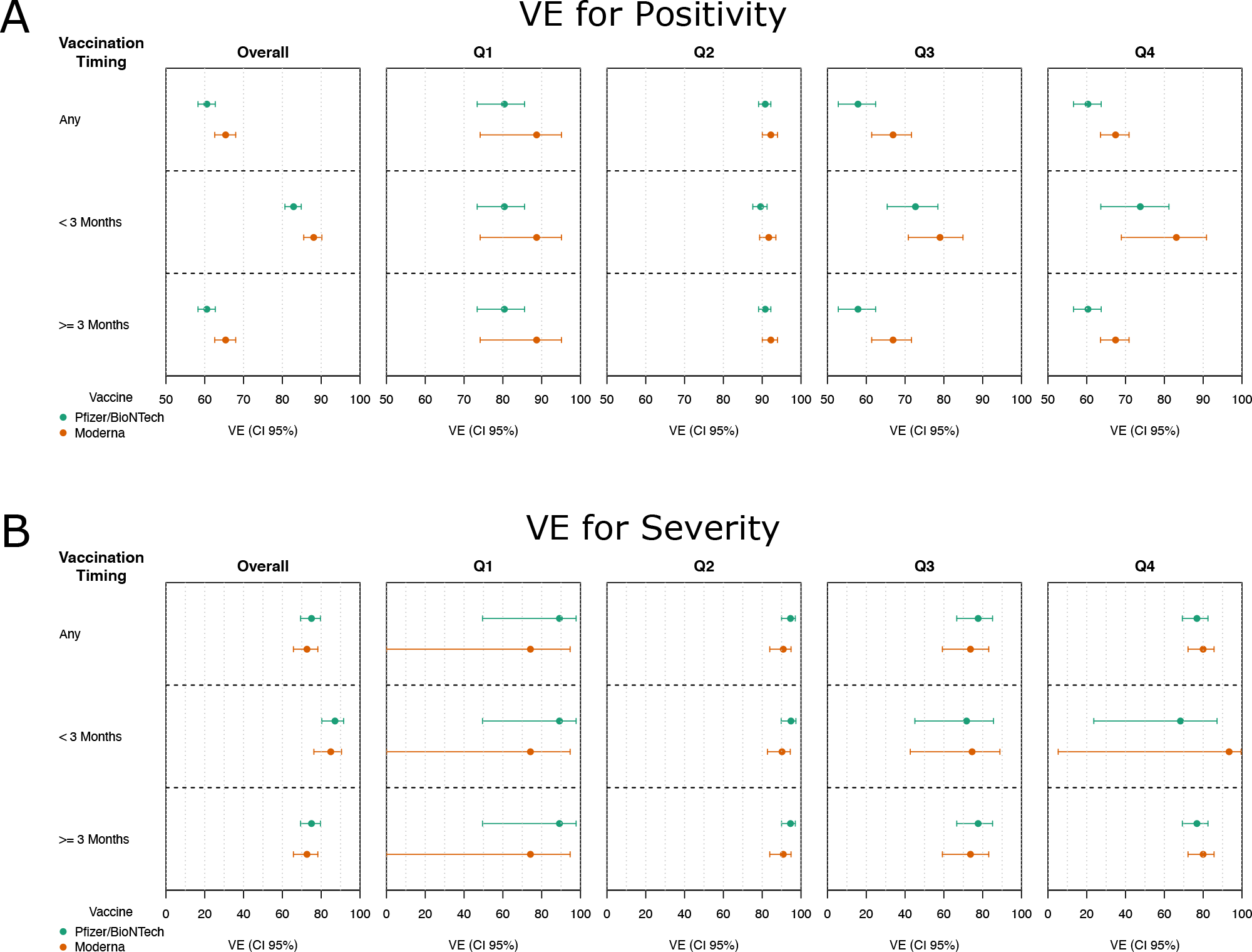
Comparison of effectiveness between Pfizer-BioNTech and Moderna vaccines. VE for Positivity (A) and VE for Severity (B) are shown for individuals being fully vaccinated at any time (top), within 3 months (center), or over 3 months (bottom) before being tested for COVID-19.

In the overall cohort, we observed lower VE-Susceptibility for Pfizer-BioNTech compared to Moderna (82.9% [80.7–84.9%] versus 88.1% [85.5–90.2%]), a trend that was consistent across the three quarters though lowest in Q2 (89.6% [87.6–91.3%] versus 91.7% [89.3–93.5%]; **Figure 2, Figure S3**). Contrarily, VE-Severity was higher for Pfizer-BioNTech (87.1% [80.3– 91.6%]) compared to Moderna (84.9% [76.2–90.5%]) in the overall cohort, consistent in the first half of 2021 but reversed in the last half of 2021. Of note, the VE-Severity in Q4 was substantially higher for Moderna (93.4% [5.3–99.6%]) compared to Pfizer-BioNTech (68.3% [23.6–87.2%]), though sample sizes were small, resulting in extremely wide and overlapping confidence intervals (**Figure S4**).

In general, we observed overlapping confidence intervals for all pairwise comparisons, suggesting similar protection against susceptibility and severity, though larger studies are needed to tease out the vaccines’ performance.

### Factors Affecting Vaccine Effectiveness

Finally, we explored if VE varied across demographic and clinical risk factors. Overall, we found that a full vaccination was moderately effective against an infection across all strata (57.1% <= VE-Susceptibility <= 65.2%) except for people who had a documented past COVID-19 infection (VE-Susceptibility _Past COVID-19 Infection_ = 47.6% [24.5% – 63.9%]; P_Int._ = 0.065); **Figure 3**). These lower estimates are expected, because unvaccinated individuals who already acquired immunity through a previous COVID-19 infection likely do not benefit as much from a vaccination as people who were not previously infected. It is important to note that only 182 individuals were reported as being reinfected in the data, and only 48 of these were fully vaccinated.

**Figure 3.**
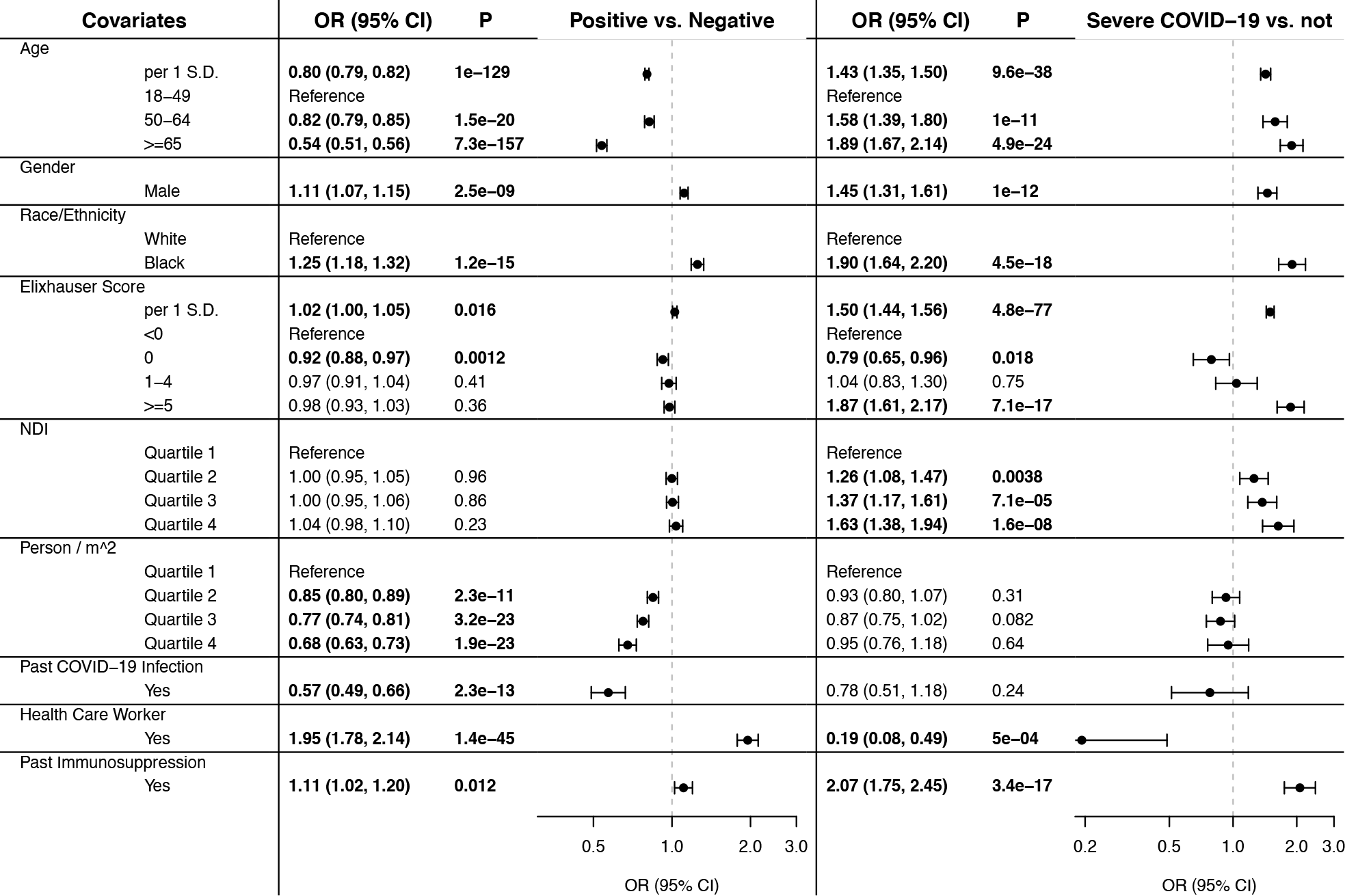
Vaccine Effectiveness across various covariate strata. Analyses were adjusted for the following set of covariates (after excluding the variable of interest): age, gender, race/ethnicity, Elixhauser Score, Persons/m^2^, NDI, Past COVID-19 Infection, and Health Care Worker Status

For health care workers (HCW) we observed a higher effectiveness compared to others (VE-Positivity _HCW_: 70.4% [62.8–76.5%] versus VE-Susceptibility _Non-HCW_: 61.7% [59.9–63.9%]; P_Int._ = 0.031), potentially indicating a stricter adherence to the vaccination regiment or additional protective measures ^20,21^.

Additionally, we observed decreased protection against an infection in individuals living in neighborhoods with increasing NDI quartiles compared to the lowest NDI quartile indicating potential health disparities (P_Int._ <= 0.023).

When we next focus on VE-Severity, we also observed effective protection against severe COVID-19 across all strata (VE-Severity >= 66.6%), except for the estimated reduced effectiveness for individuals with a past COVID-19 infection (VE-Severity _Past COVID-19 Infection_ 45.3% [0% - 77.1%]; P_Int._ = 0.133). Due to the lower counts of individuals with severe COVID-19, confidence intervals were wider (**Figure 3**). None of the observed interactions remained significant after correction for multiple testing (16 tests).

## Discussion

This study of 170,487 adults tested/diagnosed for COVID-19 at MM provides additional evidence of high COVID-19 vaccine effectiveness against SARS-CoV-2 variants across 2021. In our study, fully vaccinated individuals were more likely to test negative and were less likely to have severe disease than unvaccinated individuals or those with unknown vaccination status. Qualitatively, we observed waning effects of VE against positivity and disease severity across the four quarters. However, as the number of individuals who received booster shots increased in the fourth quarter of 2021, both infection and severity rates were lowest among boosted individuals. The Q4 increase in VE indicates improved protection through a booster when facing potential waning vaccine effectiveness and virus variants. The availability of potential confounding patient characteristics allowed us to evaluate potential modification of VE by patient subgroups. Further, our analysis compared the effectiveness of the first two FDA-approved mRNA vaccines. When including all study subjects, the Pfizer vaccine seemed to offer more protection against severe disease, while Moderna seemed to protect better against contracting COVID-19. Because Pfizer and Moderna vaccines were approved at different dates, we stratified by time since vaccination. When restricting to subsets of individuals who received the vaccine within the past three months, VE against infection tended to be slightly higher for those who received the Moderna vaccine. VE against severe disease was moderately high for both vaccines. The interpretation of changing VE is difficult as it can be affected by lower protection against the Delta or Omicron variants, waning vaccine effectiveness, or other considerations ^4^.

Our results are largely consistent with previously published findings, though differences among studies make direct comparison challenging, e.g., some studies adjusted for potential confounders (age only, other patient characteristics, or time since vaccination) while others characterized the variant (either Alpha, Mu, or Delta) associated with each case via whole-genome sequencing. Our study spans the whole year of 2021 and thus likely includes data on multiple variants (including the Omicron variant) and also on booster doses that have become more prevalent. From other studies conducted in late 2020 to early 2021, VE estimates against hospitalization ranged around 87% (CI 55-100%)^2^ and 89% (CI 87-91%)^3^ against infection. Studies during mid-2021 reported VE of 79.8%^7^ and 79.9-98.4%^4^ against infection and 95.3%^7^, 86% (CI 82-88%)^5^, 80-97.5%^4^, and 80.0% (CI 76.8-82.7%)^9^ against hospitalization across different variants. A recent study that looked further at the waning VE from late 2020 through Q3 of 2021 found VE rates for mRNA vaccines to be the highest two weeks after a two-dose vaccination regimen (94.5%; CI 94.1-94.9%), lining up with our estimates of VE in Q2, and lowest at 66.6% (CI 65.2-67.8%) after seven months^8^, which corresponds with our VE estimates in Q3. Taken together, these results highlight the need for appropriate adjustment for patient covariates, timing of vaccination, booster status, and for replication across different regions and time periods to more fully understand VE and potential VE modifying factors.

A strength of the present study is the availability of detailed EHR data for study subjects. This allows us to adjust for potential confounding patient characteristics including socioeconomic and demographic characteristics and relevant medical conditions including previous COVID-19 testing results and receipt of a booster dose. Most often, the test negative design has been used to evaluate VE^3,22,23^. As such, a limitation of results from this study design is the generalizability to individuals tested for COVID-19. Still, these studies provide insight about the long-term effectiveness and durability of vaccine protection^23^. A potential weakness is any missing information from the EHR, e.g., undocumented test results or vaccinations, though we believe categorizing individuals into an unknown/unvaccinated status tends to produce conservative VE estimates. Overall, these results should give confidence in the effectiveness of the vaccines and may encourage those who have not been vaccinated or not received a booster to do so.

## Data Availability

The datasets generated during and/or analyzed during the current study are not publicly available due to MM privacy regulations.

