## Supplemental Materials for "Estimating COVID-19 Vaccination Effectiveness Using Electronic Health Records of an Academic Medical Center in Michigan"

**Table S1** Observed series of vaccinations. 254 Individuals who received vaccines other than Pfizer/BioNTech, Moderna, or Janssen, or had an unspecified vaccine were excluded. For individuals who were tested across multiple quarters, the vaccination series at their last observed quarter are shown.

| **Vaccine / Series** | **# Vaccinations** | **n** | **Vaccine / Series** | **# Vaccinations** | **n** |
| --- | --- | --- | --- | --- | --- |
| Pfizer | 1 | 6164 | Pfizer, Pfizer, Pfizer, Pfizer | 4 | 71 |
| Moderna |  | 5399 | Moderna, Moderna, Moderna, Moderna |  | 29 |
| Janssen |  | 3773 | Pfizer, Pfizer, Moderna, Moderna |  | 9 |
| Pfizer, Pfizer | 2 | 42030 | Pfizer, Pfizer, Pfizer, Moderna |  | 6 |
| Moderna, Moderna |  | 24676 | Moderna, Moderna, Pfizer, Pfizer |  | 3 |
| Janssen, Moderna |  | 126 | Moderna, Pfizer, Moderna, Moderna |  | 2 |
| Janssen, Pfizer |  | 89 | Moderna, Pfizer, Moderna, Pfizer |  | 2 |
| Janssen, Janssen |  | 85 | Moderna, Pfizer, Pfizer, Pfizer |  | 2 |
| Moderna, Pfizer |  | 26 | Pfizer, Pfizer, Moderna, Pfizer |  | 2 |
| Pfizer, Moderna |  | 17 | Janssen, Pfizer, Pfizer, Janssen |  | 1 |
| Pfizer, Janssen |  | 7 | Janssen, Pfizer, Pfizer, Pfizer |  | 1 |
| Moderna, Janssen |  | 4 | Moderna, Moderna, Janssen, Moderna |  | 1 |
| Pfizer, Pfizer, Pfizer | 3 | 6556 | Moderna, Moderna, Moderna, Pfizer |  | 1 |
| Moderna, Moderna, Moderna |  | 2909 | Moderna, Moderna, Pfizer, Moderna |  | 1 |
| Pfizer, Pfizer, Moderna |  | 281 | Moderna, Pfizer, Pfizer, Moderna |  | 1 |
| Moderna, Moderna, Pfizer |  | 139 | Pfizer, Moderna, Pfizer, Moderna |  | 1 |
| Moderna, Pfizer, Pfizer |  | 10 | Pfizer, Moderna, Pfizer, Pfizer |  | 1 |
| Moderna, Pfizer, Moderna |  | 9 | Pfizer, Pfizer, Janssen, Pfizer |  | 1 |
| Pfizer, Pfizer, Janssen |  | 8 | Pfizer, Pfizer, Pfizer, Pfizer, Pfizer | 5 | 8 |
| Janssen, Pfizer, Pfizer |  | 7 | Moderna, Moderna, Moderna, Moderna, Moderna |  | 2 |
| Janssen, Moderna, Moderna |  | 6 | Moderna, Moderna, Moderna, Moderna, Pfizer |  | 1 |
| Pfizer, Moderna, Pfizer |  | 6 | Moderna, Pfizer, Moderna, Pfizer, Pfizer |  | 1 |
| Pfizer, Moderna, Moderna |  | 4 | Pfizer, Pfizer, Moderna, Moderna, Moderna |  | 1 |
| Moderna, Moderna, Janssen |  | 3 | Pfizer, Pfizer, Pfizer, Pfizer, Moderna |  | 1 |
| Janssen, Janssen, Janssen |  | 1 | Pfizer, Pfizer, Pfizer, Pfizer, Pfizer, Pfizer | 6 | 1 |

**Table S2. Missingness of the Variables by Vaccination Status**

| **Variable** | **Missingness, n (%)** | | | |
| --- | --- | --- | --- | --- |
|  | **Unvaccinated / Unknown (n = 78002)** | **Partially Vaccinated  (n = 18425)** | **Fully Vaccinated (n = 74060)** | **Fully Vaccinated and >= 1 Booster (n = 7187)** |
| Age | 0 (0) | 0 (0) | 0 (0) | 0 (0) |
| Gender | 8 (< 0.01) | 2 (< 0.01) | 5 (< 0.01) | 0 (0) |
| BMI | 10689 (13.7) | 1919 (10.4) | 6615 (8.9) | 369 (5.1) |
| Race Ethnicity | 0 (0) | 0 (0) | 0 (0) | 0 (0) |
| Primary Care at MM | 0 (0) | 0 (0) | 0 (0) | 0 (0) |
| Persons Per Square Mile | 14922 (19.1) | 2648 (14.4) | 12731 (17.2) | 1069 (14.9) |
| NDI without Proportion Black | 14922 (19.1) | 2648 (14.4) | 12731 (17.2) | 1069 (14.9) |
| PastCOVID-19 Infection | 0 (0) | 0 (0) | 0 (0) | 0 (0) |
| Comorbidity Score | 12849 (16.5) | 2137 (11.6) | 8351 (11.3) | 520 (7.2) |
| Elixhauser Score_AHRQ | 11777 (15.1) | 1848 (10) | 6990 (9.4) | 433 (6) |
| Health Care Worker | 0 (0) | 0 (0) | 0 (0) | 0 (0) |
| Past Immunosuppression | 0 (0) | 0 (0) | 0 (0) | 0 (0) |

**Table S3.** Settings and nested sets of covariate adjustments

| **Covariate** | **Data Type** | **FET*** | **Firth’s bias adjusted logistic regression** | | | | | |
| --- | --- | --- | --- | --- | --- | --- | --- | --- |
|  |  | **Unadjusted** | **Unadjusted** | **Adj. 1** | **Adj.2** | **Adj.3** | **Adj.4** | **Adj.5** |
| Age | Continuous | … | … | Included | Included | Included | Included | Included |
| Gender | Binary | … | … | Included | Included | Included | Included | Included |
| Self-reported  Race / Ethnicity | Categorical (4 levels**) | … | … | Included | Included | Included | Included | Included |
| Elixhauser Score | Continuous | … | … | … | Included | Included | Included | Included |
| NDI without proportion black | Categorical (quartiles) | … | … | … | … | Included | Included | Included |
| Population density | Categorical (quartiles) | … | … | … | … | Included | Included | Included |
| Past COVID-19 Infection | Binary | … | … | … | … | … | Included | Included |
| Health Care Worker Status | Binary | … | … | … | … | … | … | Included |

* Fisher's Exact Test for Count Data

** Self-reported Race / Ethnicity factor levels: Caucasian/Non-Hispanic, African American/Non-Hispanic, Other Race or Ethnicity, and Unknown Race or Ethnicity

**Figure S1** Comparison of effectiveness for those who were fully vaccinated with and without a booster shot. VE Susceptibility and VE Severity are shown for individuals stratified by booster status in Q4. VE was estimated using logistic regression adjusting for Age, Gender, Race/Ethnicity, Elixhauser Score AHRQ, Persons Per Square Mile, NDI without Proportion Black, Past COVID-19 Infection and Health Care Worker Status (Adjustment 5)

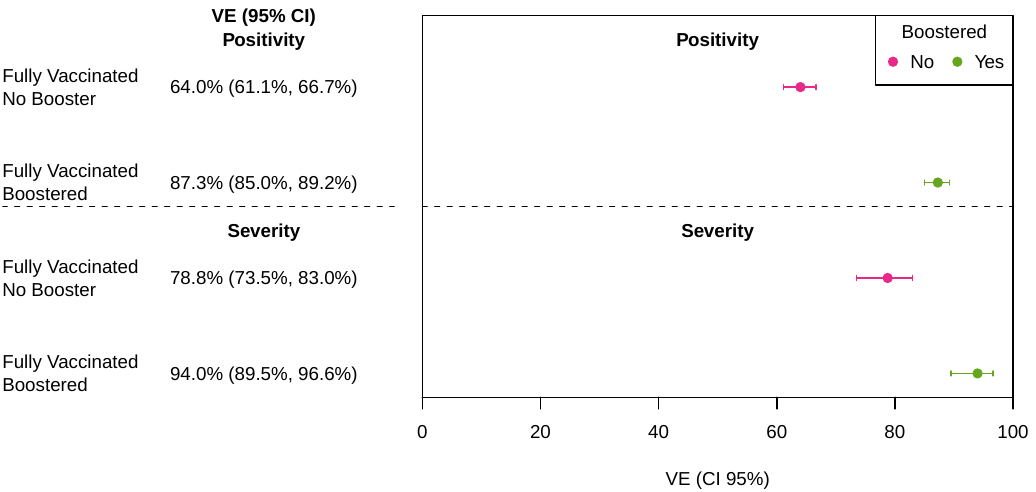

**Figure S2**. Vaccine Effectiveness in individuals who received primary care at MM. (A) Positivity and (B) Severity

A

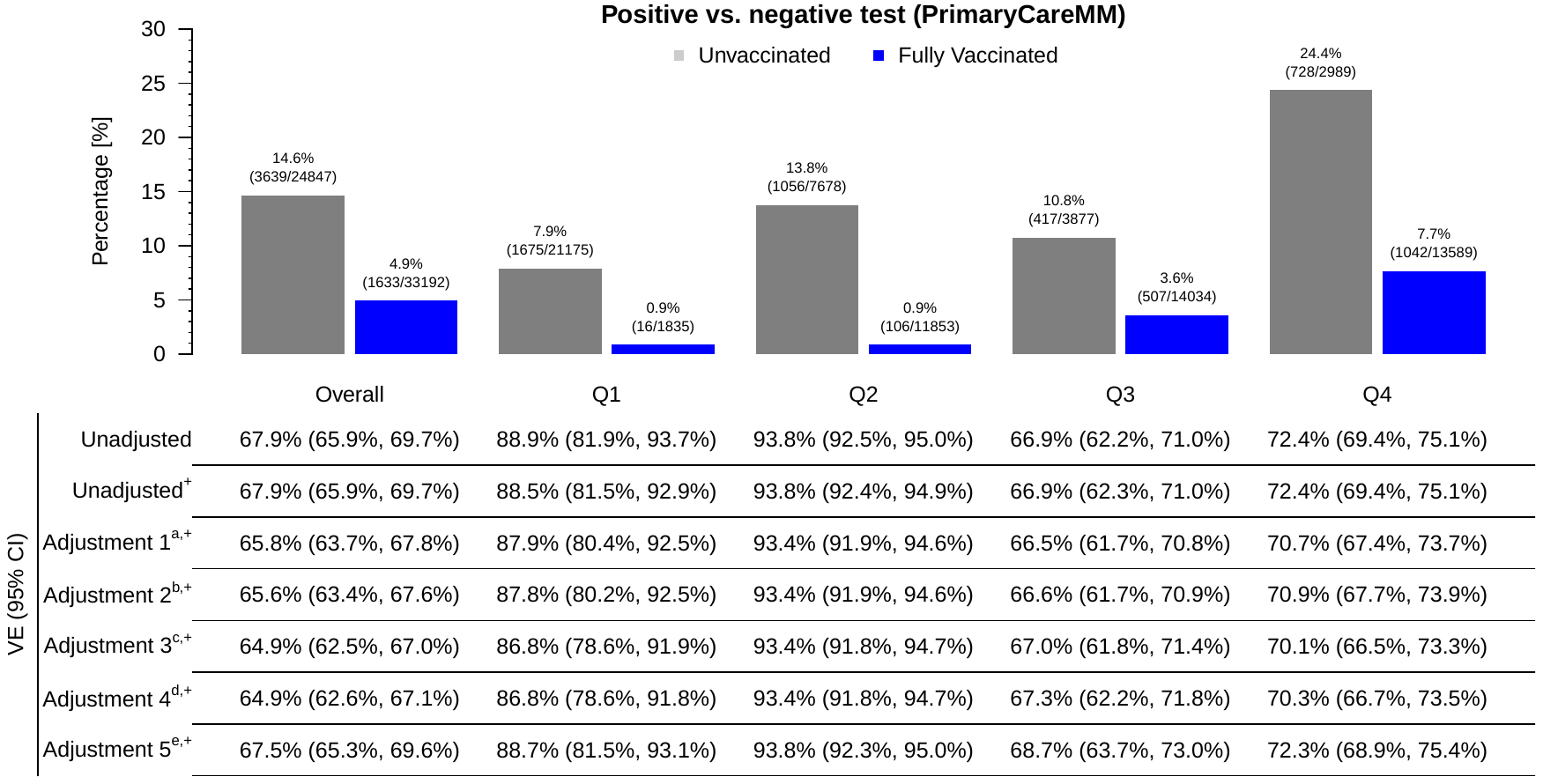

B

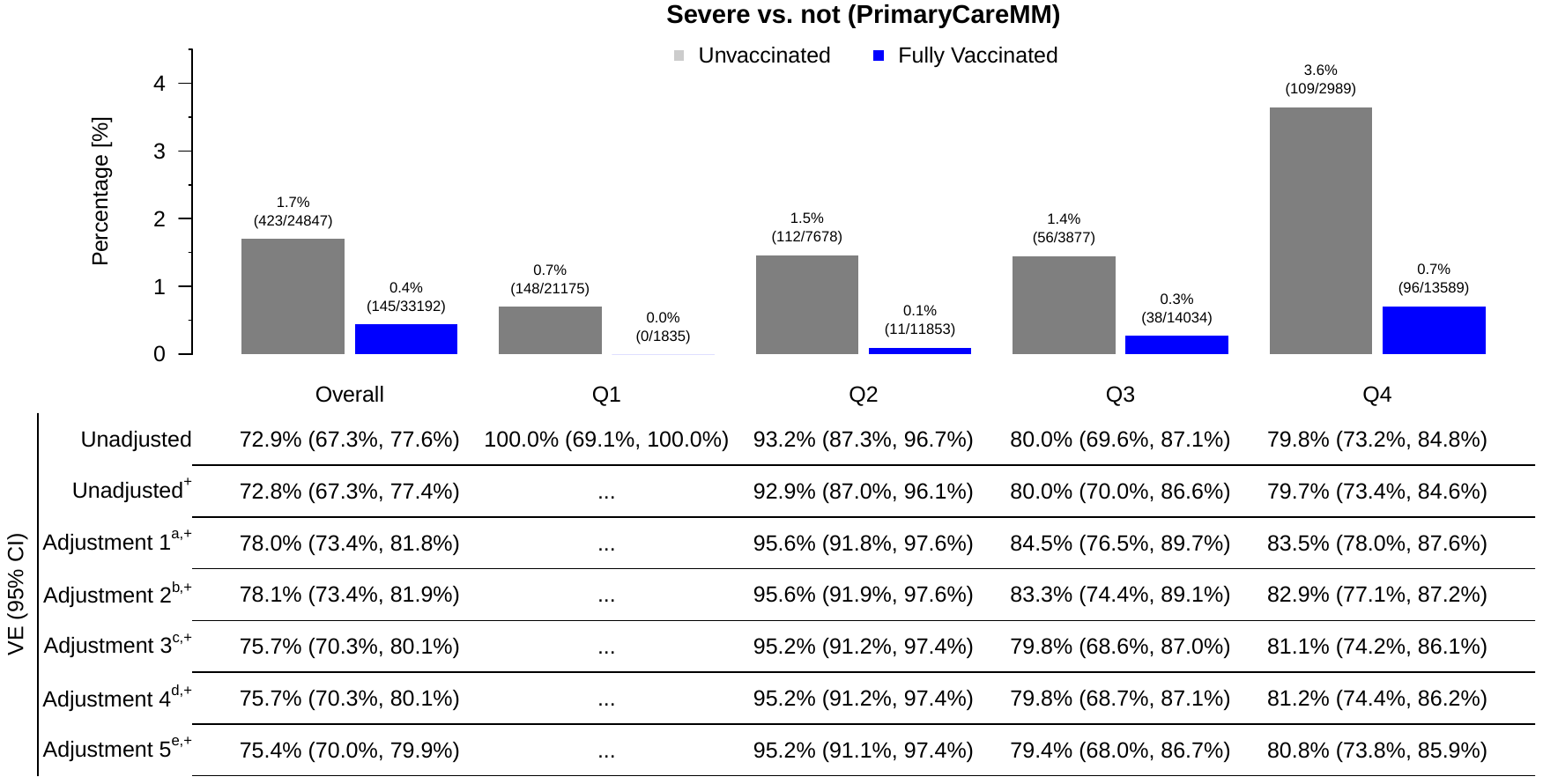

+: Logistic regression; a: Age, Gender, Race/Ethnicity; b: Adjustment 1 + Elixhauser Score AHRQ; c: Adjustment 2 + Persons Per Square Mile + NDI without Proportion Black; d: Adjustment 3 + Past COVID-19 Infection; e: Adjustment 4 + Health Care Worker Status

**Figure S3.** Vaccine effectiveness (positivity) in individuals who received (A) Pfizer-BioNTech or (B) Moderna. Only individuals who were consider fully vaccinated less than 3 months before their test or diagnosis and did not receive an additional booster shot were included.

A

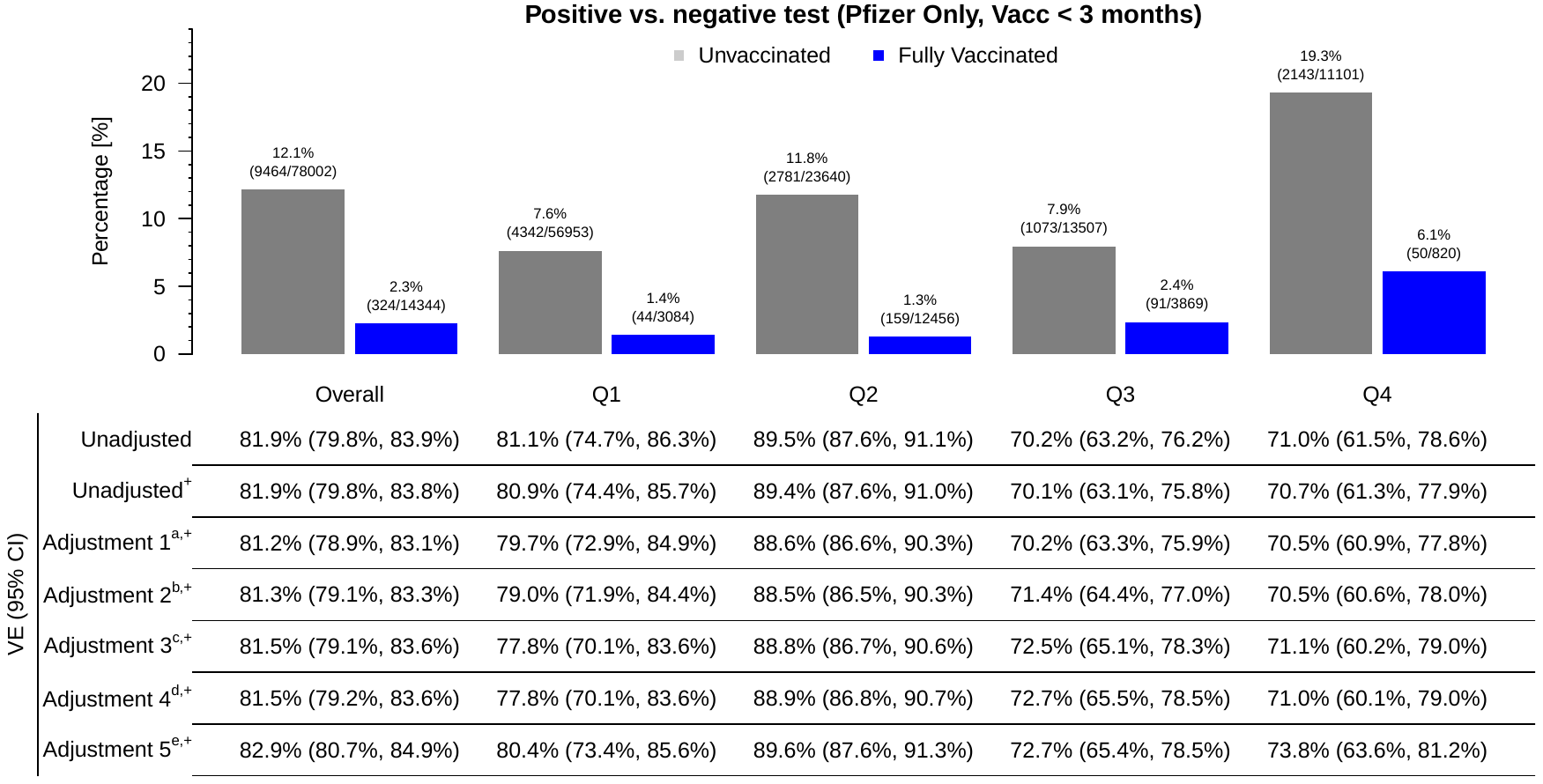

B

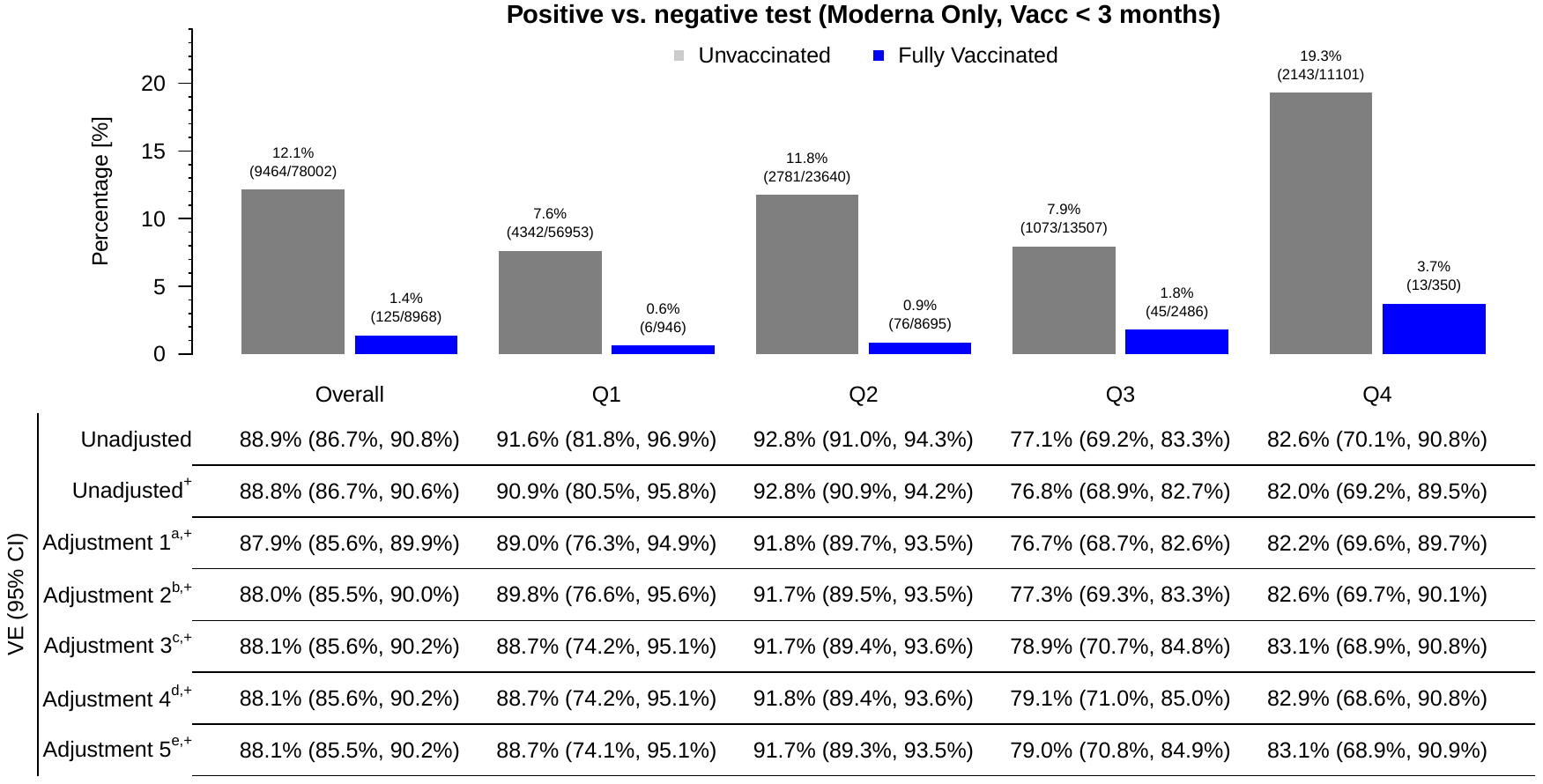

+: Logistic regression; a: Age, Gender, Race/Ethnicity; b: Adjustment 1 + Elixhauser Score AHRQ; c: Adjustment 2 + Persons Per Square Mile + NDI without Proportion Black; d: Adjustment 3 + Past COVID-19 Infection; e: Adjustment 4 + Health Care Worker Status

**Figure S4.** Vaccine effectiveness (severity) in individuals who received (A) Pfizer-BioNTech or (B) Moderna. Only individuals who were consider fully vaccinated less than 3 months before their test or diagnosis and did not receive an additional booster shot were included.

A

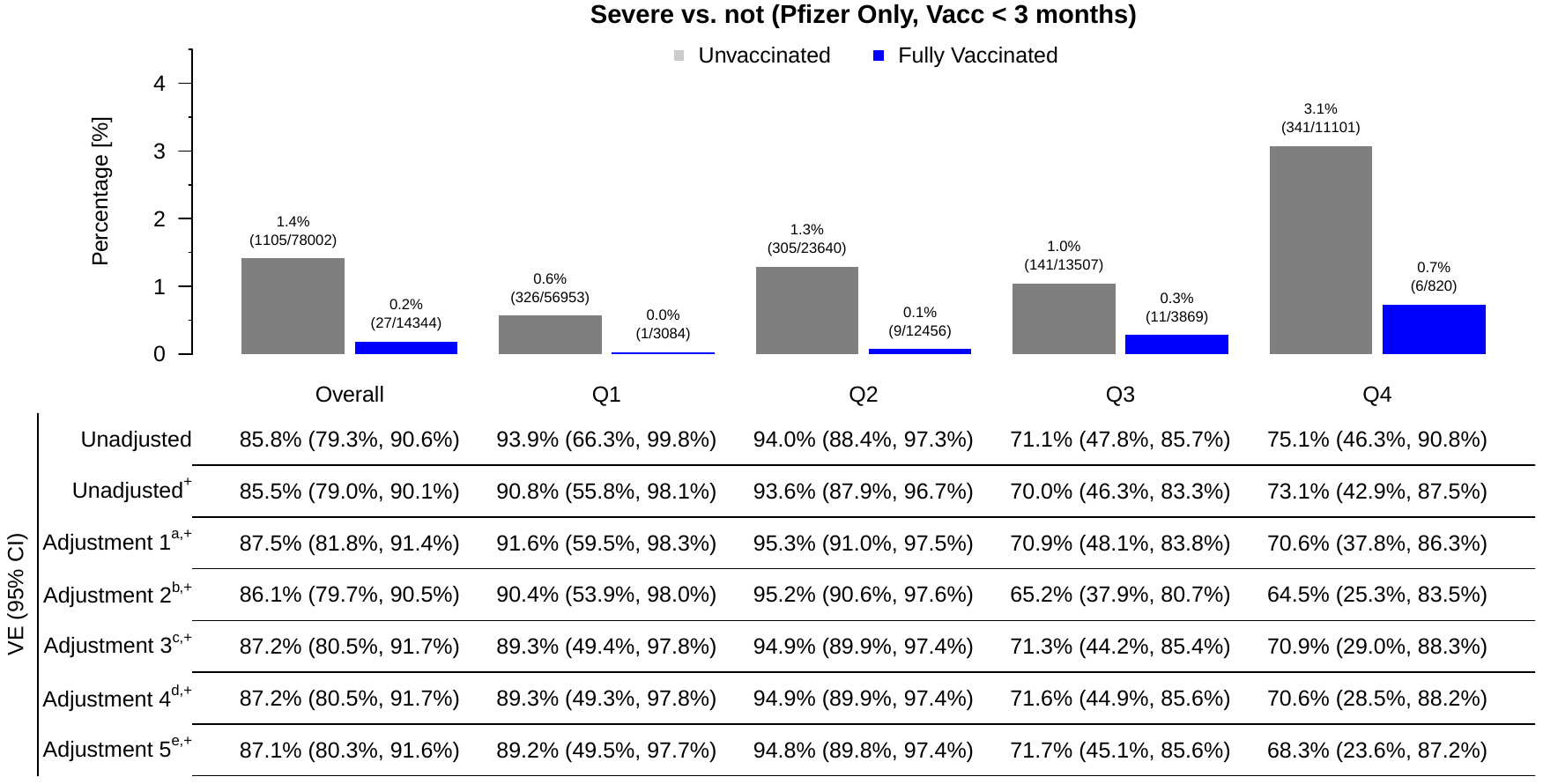

B

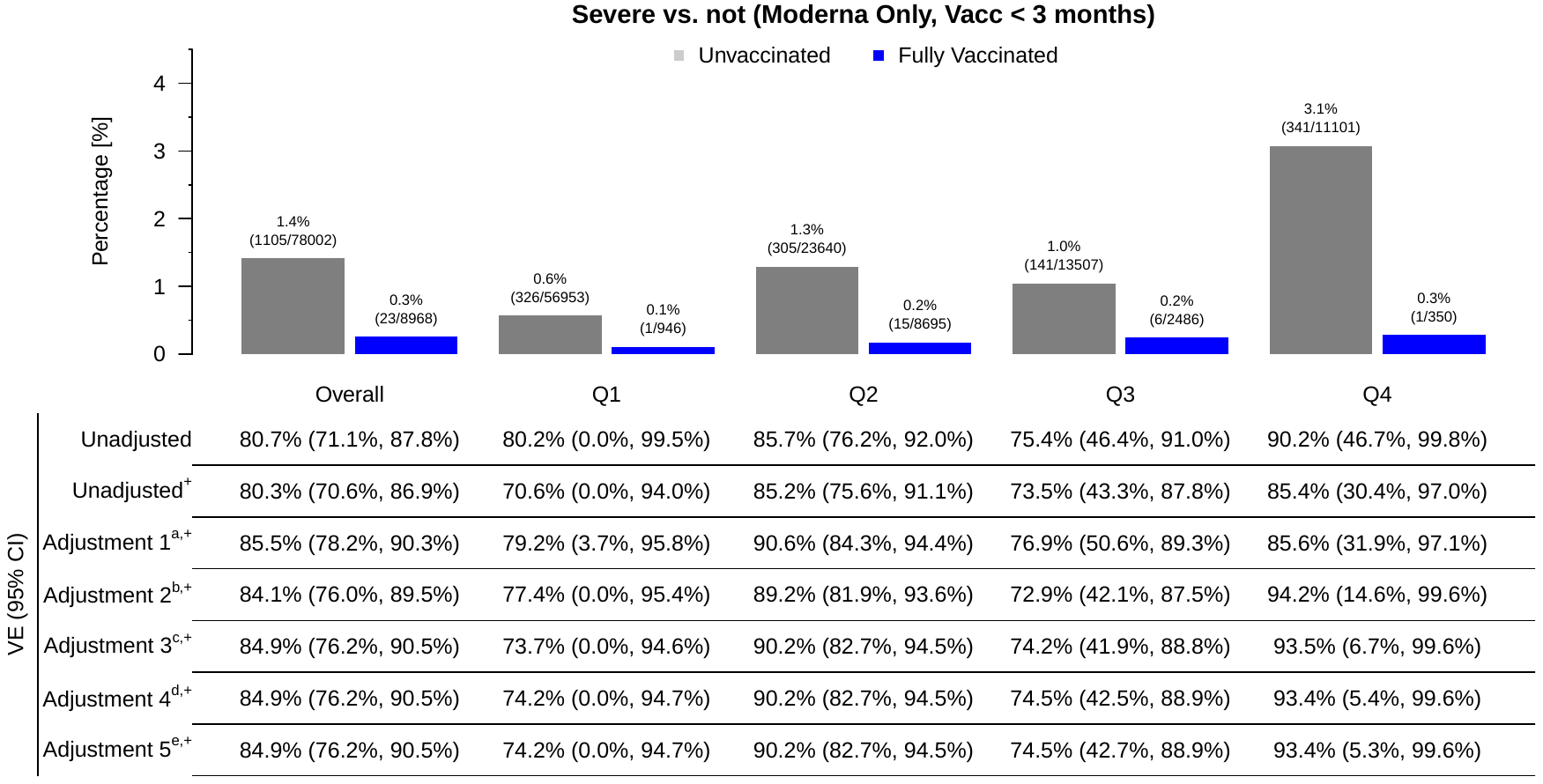

+: Logistic regression; a: Age, Gender, Race/Ethnicity; b: Adjustment 1 + Elixhauser Score AHRQ; c: Adjustment 2 + Persons Per Square Mile + NDI without Proportion Black; d: Adjustment 3 + Past COVID-19 Infection; e: Adjustment 4 + Health Care Worker Status
